# Off-label use in Palliative Care – more common than expected. A retrospective chart review

**DOI:** 10.1101/2021.01.17.21249962

**Authors:** Vera Hagemann, Claudia Bausewein, Constanze Rémi

## Abstract

**Objective:** Off-label drug use seems to be integral to palliative care pharmacotherapy. Balancing potential risks and benefits in the context of limited therapeutic options is challenging. To provide specific support for clinicians in dealing with off-label use, it is essential to understand off-label use in everyday clinical practice.

The aim of this pilot study was to quantify and describe off-label use in a palliative care unit.

**Methods:** Retrospective chart review of all adult patients treated on a palliative care unit in 10/2017. All data on drug use e.g. indication, dose, route of administration were extracted and matched with the prescribing information. Identified off-label use was subsequently compared with recommendations in the relevant literature. The main outcome measure was frequency and type of off-label drug use.

**Results:** 2,352 drug application days (d) and 93 drugs were identified for 28 patients. Of all drugs, 47 (51%) were used off-label at least once. Most off-label uses concerned indication (57%), followed by mode of administration. In drugs highly relevant to palliative care the rate of off-label use was as high as 67%. The extent to which off-label therapy was supported by literature was very variable and ranged from 0 to 88%.

**Conclusions:** This single-unit data confirms the high prevalence of off-label use in palliative medicine and demonstrates that off-label use in palliative care is very multifaceted. The data presented allows for a more precise characterization of various aspects of off-label use in order to derive concrete further measures for research and clinical practice.

**What is already known on this subject:**

- Off-label drug use is likely to be common in palliative care, but detailed data is very limited
- Off-label drug use is a potential threat for patient safety
- Physicians state to make therapeutic decisions based on their own experience, due to a lack of available evidence and lack of support in assessment

**What this study adds:**

- off-label use in palliative care is multifaceted
- the mode of administration (e.g. combination with other drugs in a syringe driver) is beside indication a common reasons for off label use
- the proportion of off-label use without sound evidence is high.

## Introduction

The aim of palliative care is to improve quality of life of patients with advanced diseases. Drug therapy is a mainstay in the treatment of physical symptoms. However, based on the few data available so far, it can be assumed that up to a quarter of all prescribed drugs are used outside the scope of the marketing authorization in palliative care^1-4^. In general, “off-label use” refers to all deviations from the approval (license) of the drug, for example with regard to indication, route of administration, dosage interval or duration of treatment^5, 6^.

In contrast to other medical disciplines, the management of patients towards the end of life does not focus on curative or disease-modifying treatment approaches but on the alleviation of distressing symptoms. Often, only the route of administration or dosage differs from the manufacturer’s approval. The existing evidence for off-label use in palliative medicine is very heterogeneous, e.g. with good evidence for the use of opioids for breathlessness^7^ but only very limited evidence with case reports and small studies on the frequent practice of subcutaneous infusion of midazolam (e.g.^8, 9^).

Off-label use involves both medical and legal challenges. It always carries the potential risk of a drug that has not been tested or insufficiently tested for its intended use and thus is a potential threat for patient safety^10^. Additionally, off-label use can lead to limited coverage of treatment costs by insurance companies or changes in liability for treatment related harm^11^.

Accordingly, it should be applied consciously and well-considered. In clinical practice, however, there is often a lack of time and resources to conduct a patient-specific risk-benefit analysis for each therapy on the basis of the current literature and available alternatives^1^. Achieving the Hippocratic principle of “Primum non nocere” (“first, do no harm”) is therefore often a major challenge. Physicians regularly make therapeutic decisions based on their own experience, especially due to lack of available evidence and lack of support in assessment^11^. They express concerns about the safety of drug therapy^11^. To date, there are only limited data to provide a sound evidence-base for decision-making on the use of drugs in palliative medicine. In addition, such studies are difficult to conduct due to methodological difficulties. Pharmacists can play an important role in the off-label medication process. Potential tasks include: identification of off-label therapies, assisting in the interpretation of the available evidence, evaluation of possible treatment alternatives and support in the patient-specific benefit-risk assessment^12^.

For providing specific support for clinicians in dealing with off-label use, it is essential to understand off-label use in everyday clinical practice. This includes not only the prevalence but also the clinical circumstances, the underlying evidence and the decision-making processes. Based on this knowledge it is only possible to decide whether measures should be taken to increase safety of therapy and make it possibly more effective for the patient. At the same time, however, such measures can also help to give prescribers more security in dealing with off-label use.

As little is known about off-label use in palliative medicine in Germany and internationally^1, 11^, the aim of this pilot study was to quantify and describe off-label use in adult patients treated on a palliative care unit.

## Methods

Medical charts of patients treated in the palliative care unit of the Department of Palliative Medicine at Munich University Hospital between 1^st^ and 31^st^ October 2017 were retrospectively reviewed. The reporting of this chart review complies with the STROBE criteria for reporting cross-sectional studies^13^.

The palliative care unit at Munich University Hospital provides acute palliative care for patients with advanced disease. Annually, about 300 patients are treated in the unit (286 in 2017) of which about 40% are discharged. Patients suffer predominantly from malignant (about 75%) and non-malignant disease.

Prescribing data was extracted for all drugs administered on the unit during October 2017 using an extraction sheet. This month was randomly selected from all months of the year with typical patient case numbers for the unit. The extraction sheet was piloted by two persons and subsequently modified. Data was extracted by two pharmacy students and data integrity randomly checked by two pharmacists. The recording encompassed active generic drug, trade name, indication, dose, dosing interval, route of administration, mode of administration, and duration of therapy. Drug applications were recorded as application days, i.e. every day on which a drug was administered per patient and not every single dose was counted. This was to avoid bias due to short rather than long dosing intervals. The data obtained was compared with the prescribing information to identify off-label applications. An application day was rated as off-label when a drug was applied for an aspect outside the scope of the marketing authorisation regarding indication, drug dose, dosing interval, route of administration, mode of administration (e.g. crushed, via feeding tube, combination with other drugs in syringe driver, continuous infusion), and duration of therapy (s. table 1). This was also the case if, for example, morphine was used in an approved indication (pain) but mixed with other substances in the same infusion for administration. Off-label use was subsequently compared with recommendations in the German Guideline on Palliative Medicine for Patients with Incurable Cancer^14^ and the German Palliative Care Formulary^15^ in order to identify drug uses outside the scope of the marketing authorisation but within the range of official or accepted therapy recommendations.

**Tab. 1.**
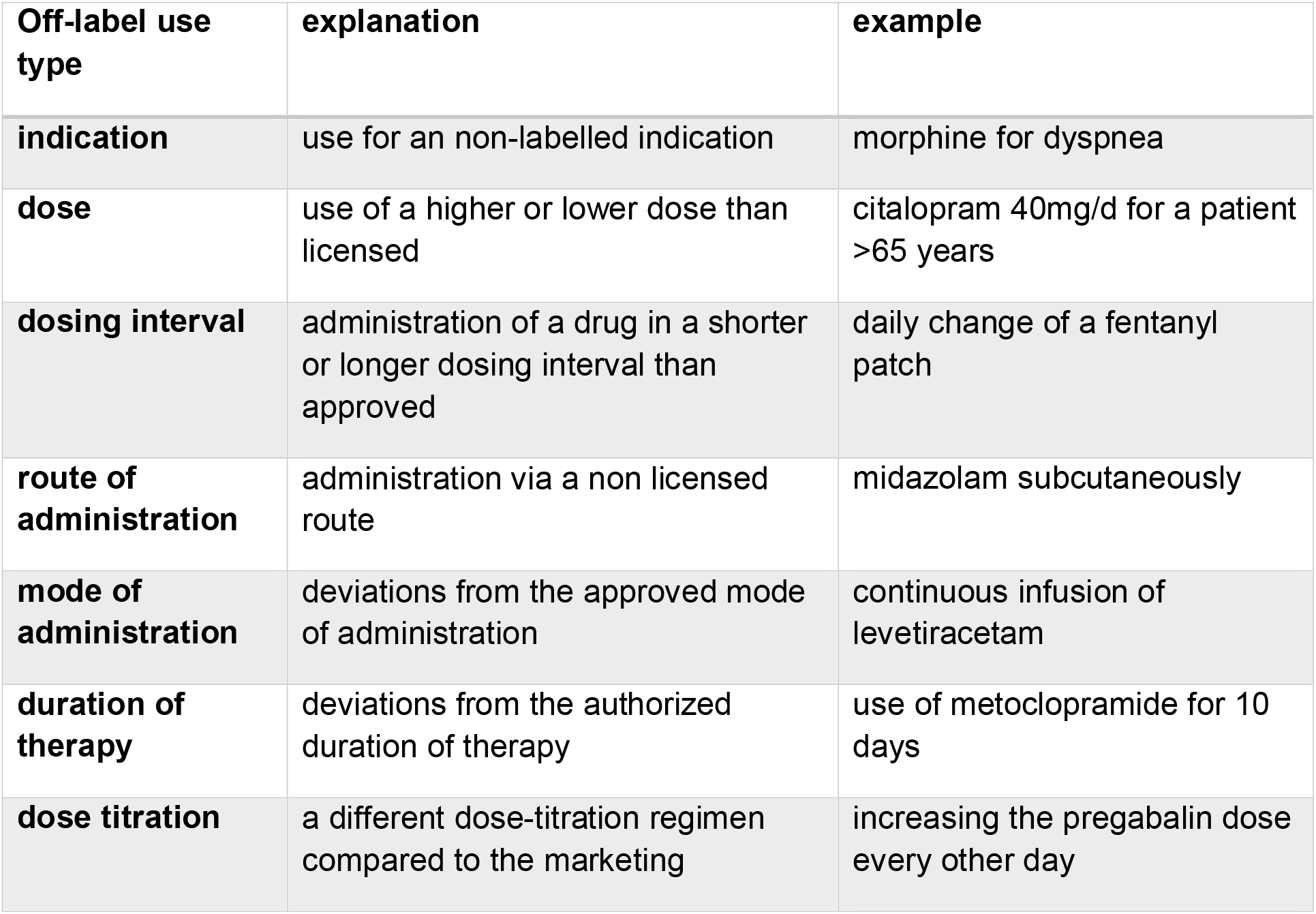
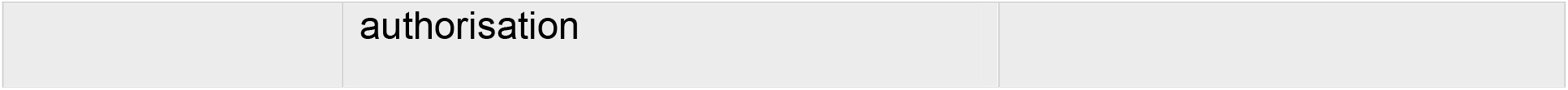
Off-label use types with explanations and examples from palliative care practice

In addition, the following patient data were recorded anonymously: age, disease (malignant/non-malignant) and sex.

The evaluation of ambiguous prescriptions was discussed between two pharmacists, for example several or unclear indications of a drug.

A committee consisting of two pharmacists and a doctor additionally divided the identified drugs into three categories based on their relevance in symptom control in palliative medicine: 1. high relevance, 2. medium relevance, 3. low relevance.

Drugs in the first group (high relevance) included those typically used in symptom control in palliative medicine based on published data and personal experience^14, 16, 17^, e.g. opioids, non-opioid analgesics, antiemetics, antipsychotics, benzodiazepines, antidepressants, antiepileptics, laxatives, anticholinergics and ketamine. Drugs with medium relevance were those used relatively regularly to prevent complications (e.g. anticoagulants, levothyroxine) but were not prescribed for distressing symptoms. In addition, drugs with medium relevance were also those used less frequently (e.g. bisphosphonates, drugs for Parkinson’s disease) or for a questionable indication, e.g. proton pump inhibitors and various drugs for insomnia.

Drugs considered to be of low relevance were generally those used for comorbidities with no or little impact on current distressing symptoms, e.g. antihypertensives, antidiabetics.

Descriptive statistics (frequencies and percentages) were calculated to describe and summarize all variables using Microsoft excel (version 1902).

The Research Ethics Committee of the Ludwig-Maximilians-Universität Munich granted ethical approval for this study (19-445).

## Results

The chart review identified a total of 2,352 drug application days (d). Over the period of one month, 93 different drugs were administered to 28 patients (15 males; 54%) with a median age of 74 years (range 41-95) on 284 patient days. All but four patients suffered from malignant disease. Of all drugs, 47 (51%) were used off-label at least once.

Of 2,352 application days, 1,248 days (53%) were off-label. Most off-label uses concerned the indication (56.5%), followed by mode of administration, drug dose, and route of administration (see Fig.1).

**Fig. 1.**
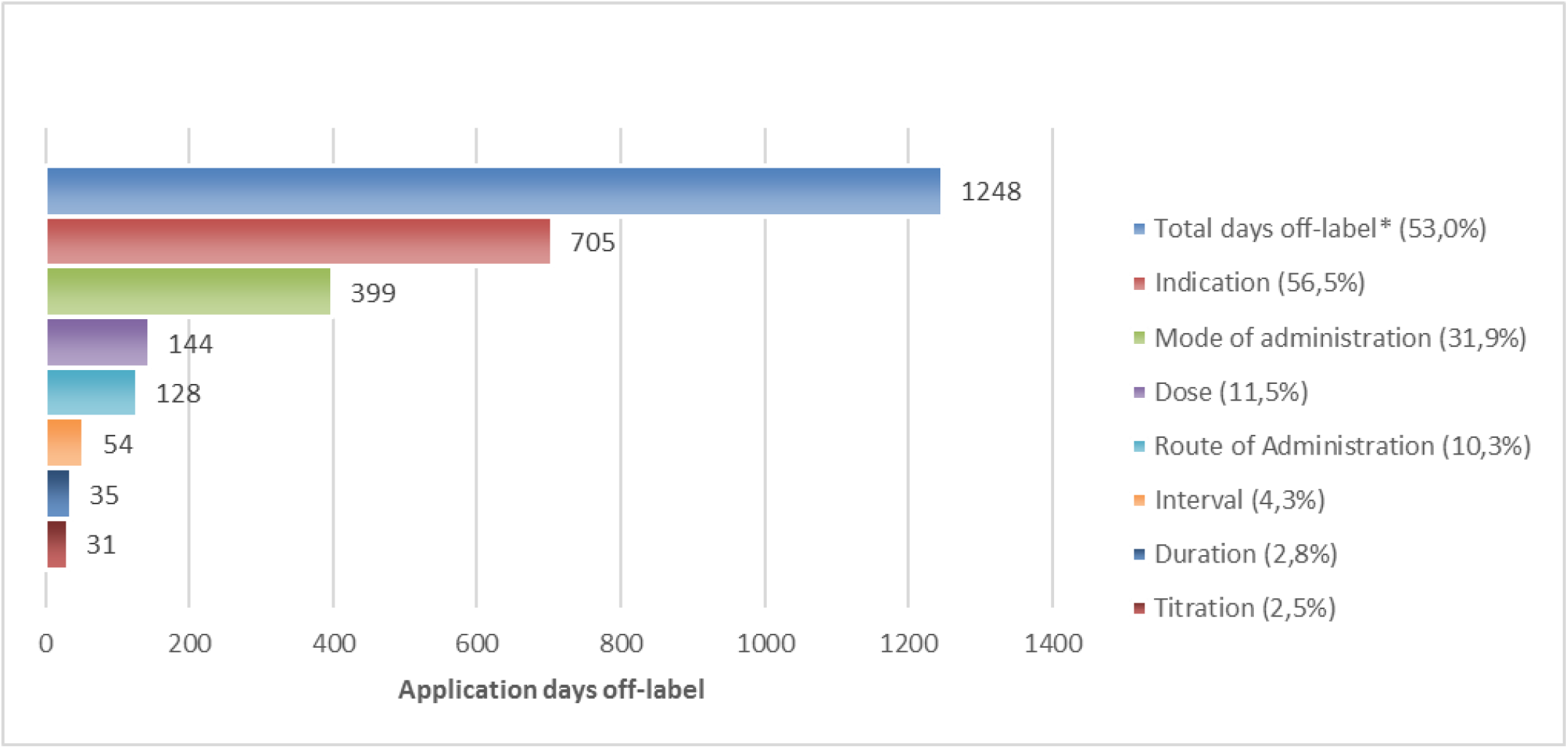
Frequency of off-label application days by type of off-label use. total number of drug applications days (in-label and off-label): 2,352 **\***off-label uses could occur in more than one category resulting in a higher total number of off-label uses than application days.

**Fig. 2.**
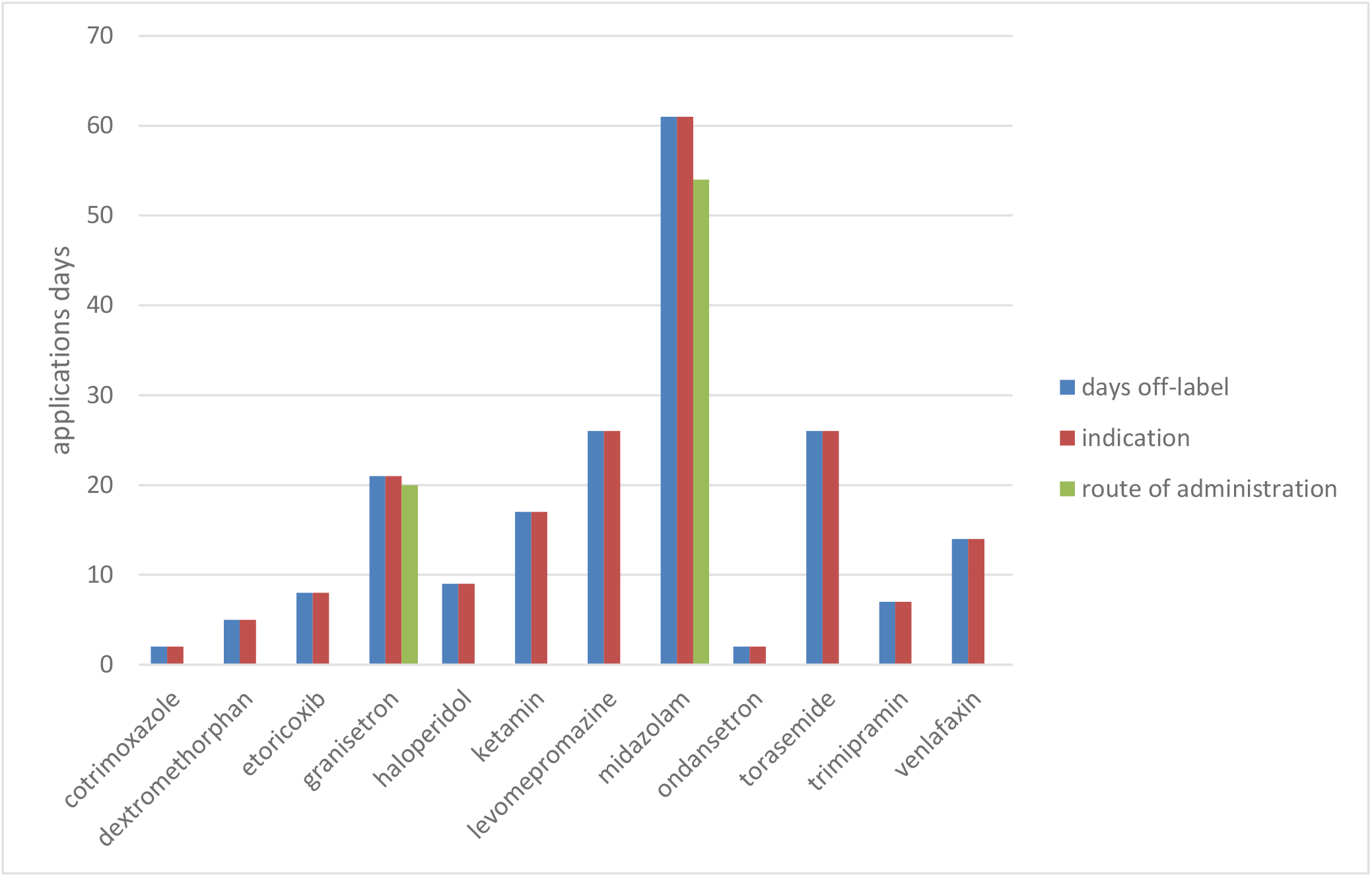
Applications days of drugs solely used off-label. Only drugs that have solely been used outside the approved indication are listed.

The drugs identified were pooled in 19 groups (Tab. 2), according to their pharmacologic group or target symptom. The grouping was decided after discussion in the research team. It should be noted that some drugs could appear in more than one group if there was more than one indication or the indication remained unclear, e.g. melperone and levomepromazine as antipsychotics and hypnotics.

**Tab. 2.** Drug groups used during the study period (descending order of off-label use frequency)

| **Drug group** | **Off-label (%)** | **Application days** | **In-label (d)** | **Off-label (d)** |
| --- | --- | --- | --- | --- |
| **Anaesthetics<sup>a</sup>** | 100.0% | 17 | 0 | 17 |
| **Antipsychotics** | 89.5% | 95 | 10 | 85 |
| **Antidepressants** | 84.3% | 153 | 24 | 129 |
| **GIT-symptoms<sup>b</sup>** | 73.1% | 186 | 50 | 136 |
| **Corticosteroids** | 71.0% | 148 | 43 | 105 |
| **Opioids** | 70.4% | 328 | 97 | 231 |
| **Benzodiazepine** | 69.8% | 96 | 29 | 67 |
| **Hypnotics** | 65.4% | 228 | 79 | 149 |
| **Antiemetics** | 63.1% | 195 | 72 | 123 |
| **Respiratory tract<sup>c</sup>** | 46.0% | 224 | 121 | 103 |
| **Diuretics** | 45.8% | 72 | 39 | 33 |
| **Anti-infectives** | 38.1% | 42 | 26 | 16 |
| **Other** | 34.8% | 178 | 116 | 62 |
| **Non-opioid analgesics** | 29.8% | 188 | 132 | 56 |
| **Anticoagulation** | 18.6% | 102 | 83 | 19 |
| **Antiepileptics** | 17.8% | 174 | 143 | 31 |
| **Laxatives** | 0% | 130 | 130 | 0 |
| **Cytostatics** | 0% | 13 | 13 | 0 |
| **Bone metabolism** | 0% | 21 | 21 | 0 |

<sup>a</sup>Ketamine only drug in group
<sup>b</sup>Drugs for gastrointestinal symptoms (not antiemetics): esomeprazole, hyoscine butylbromide, magaldrat, pantoprazole, saccharomyces boulardii, simeticon
<sup>c</sup>Drugs for respiratory tract: ambroxol, budesonide, codein, dextromethorphan, formoterol, hyoscine butylbromide, ipratropium bromide, salbutamol, tiotropium bromide, tyloxapol
<sup>d</sup>Other drugs: amlodipine, bisoprolol, epoetin, levodopa/benserazid, levothyroxine, metoprolol, naloxone, sage extract, sitagliptin, sodium bicarbonate, sodium polystyrene sulfonate

### Prevalence of off-label use in different therapeutic groups

The five groups with the most application days were opioids (328 d; 14%), hypnotics (228 d; 10%), respiratory tract (224 d; 10%), antiemetics (195 d; 8%), and non-opioid analgesics (188 d; 8%), see Tab.1. The group with the highest rate of off-label applications were anaesthetics, with ketamine as the only group representative. All 17 days of use were outside the license, as it was used exclusively in the non-approved indication of pain. A likewise large number of uses beyond the license was documented for antipsychotics (95 d; 89%). Of these, 68 application days were related to non-licensed indications (80%). The drug mainly used in this group was quetiapine with 52 application days and an off-label/in-label ratio of 96% (50 d). Of these, 33 application days (66%) were for off-label indications like delirium or depression and supported by scientific literature. Haloperidol (9d; 100%) and levomepromazine (26d; 100%) were used less frequently, but only outside the approved indication (both used for nausea).

Within the group of opioids, morphine was the drug most commonly used with 152/329 application days (46%). Of the 152 application days, 133 days were off-label (88%); 101 days (81%) were for the off-label indication dyspnea and supported by literature recommendations. The remaining 51 applications days (38%) were related to a non-licensed use in combination with other drugs in the same syringe.

Antidepressants (156 d of all application days; 7%), corticosteroids (148 d of all application days; 6%), drugs for anticoagulation (102 d of all application days, 4%), diuretics (72 d of all application days; 3%) and anaesthetics (17d of all application days; 0.7%) differed from all other groups as all uses beyond the license were solely for off-label indications and not for other reasons. In the antidepressant group, mirtazapine (112 d), trimipramine (7 d) and venlafaxin (14 d) summed up to a total of 133 days and 97% off-label/in-label-ratio. In the corticosteroid group, all off-label administrations were related to dexamethasone (105 d off-label use; 8% off-label use). Of these five drug groups, diuretics had a relatively low off-label/in-label-ratio (33 d off-label use; 46%), and the off-label days were mainly caused by the use of torasemide (26d) for a differing indication. Dexamethasone and torasemide were both prescribed for edema, dexamethasone additionally for pain. Mirtazapine and trimipramine were used for insomnia, mirtazapine additionally for pain. Venlafaxine was used for pain and aspirin in atrial fibrillation. None of these indications were supported by the literature specified above.

In addition to the drug groups, another 20 drugs (22% of all drugs used) were only used off-label. Twelve drugs were always used in differing indications, s. fig 2 Other drugs with 100% off-label application days were bisoprolol, levomethadone, meropenem, simeticone, sitagliptin, budesonide/formoterol, thyme juice, and tilidine.

### Drug relevance in palliative care

The 93 drugs used during the observation period were stratified into groups according to their relevance for palliative care pharmacotherapy (high, medium, low). 30 (32%) drugs were considered as highly relevant, e.g. ketamine, haloperidol, hyoscine butylbromide, and morphine. The relevance is also reflected by the total application days of this group. Of 1432/2352 (67%) application days attributable to drugs in this group, 902 days (63%) were off-label. For 14 of these drugs some evidence for off-label use could be identified in the literature screened. The type of off-label use and underlying evidence is displayed in table 3.

**Table 3.** Type of off-label use for drugs highly relevant in palliative care dring the study period (displayed in application days)

|  | Type of off-label use (displayed in applications days) |  |  |  |  |  |  |  |  |  |  |
| --- | --- | --- | --- | --- | --- | --- | --- | --- | --- | --- | --- |
|  | Applic<br>ation<br>days | Off-label<br>use | Indi-<br>cation | Dose | Inter-<br>val | Titra-<br>tion | Du-<br>ration | Mode<br>of<br>Ad-<br>minis-<br>tration | Route | Off-label<br>indication | Under-<br>lying<br>evi-dence |
| **Buprenorphine** | 4 | - | - | - | - | - | - | - | - | - | - |
| **Butylscopolamin** | 28 | 27 | 21 | - | - | - | - | 36 | - | mostly<br>noisy<br>secretions | German<br>Guideline,<br>APM |
| **Dexamethasone** | 134 | 105 | 105 | - | - | - | - | - | 8 | mostly<br>edema | German<br>Guideline,<br>APM |
| **Dimenhydrinate** | 12 | 6 | - | - | - | - | - | - | 6 | - | - |
| **Domperidone** | 1 | - | - | - | - | - | - | - | - | - | - |
| **Fentanyl** | 33 | 21 | 21 | - | - | - | - | - | - | dyspnea | German<br>Guideline,<br>APM |
| **Granisetron** | 21 | 21 | 18 | - | - | - | - | 2 | 20 | nausea/vo<br>miting<br>(not<br>chemother<br>apy<br>associated<br>) | APM |
| **Haloperidol** | 9 | 9 | 9 | - | - | - | - | - | 2 | nausea | APM |
| **Hydromorphone** | 75 | 49 | 22 | - | - | - | - | 25 | - | dyspnea | German<br>Guideline,<br>APM |
| **Ketamine** | 17 | 17 | 17 | - | - | - | - | 8 | - | pain | APM |
| **Lacosamide** | 23 | 5 | - | - | - | 5 | - | - | - | - | - |
| **Sodiumpico-**<br>**sulphate** | 55 | - | - | - | - | - | - | - | - | - | - |
| **Levetiracetame** | 49 | 20 | - | - | - | 3 | - | 12 | 13 | - | - |
| **Levomepromazin** | 26 | 26 | 26 | - | - | - | - | 4 | 15 | nausea/vo | APM |

Table 3. Type of off-label use for drugs highly relevant in palliative care during the 250 study period (displayed in application days)
| e |  |  |  |  |  |  |  |  |  | miting |  |
| --- | --- | --- | --- | --- | --- | --- | --- | --- | --- | --- | --- |
| Levomethadone | 1 | 1 | - | - | - | - | - | 1 | - | - | - |
| Lorazepam | 35 | 6 | - | - | 2 | - | - | 4 | - | - | - |
| Metoclopramide | 101 | 65 | - | 6 | - | - | 35 | 44 | 4 | - | - |
| Melperone | 8 | - | - | - | - | - | - | - | - | - | - |
| Metamizole<br>(dipyrone) | 174 | 48 | - | - | - | - | - | 96 | 13 | - | - |
| Midazolam | 61 | 61 | 61 | - | - | - | - | 58 | 22 | agitation,<br>anxiety | German<br>Guideline,<br>APM |
| Mirtazapine | 112 | 108 | 108 | - | - | - | - | - | - | insomnia,<br>pain | APM |
| Morphine | 152 | 133 | 101 | - | - | - | - | 51 | - | dyspnea | German<br>Guideline,<br>APM |
| Movicol | 69 | - | - | - | - | - | - | - | - | - | - |
| Ondansetron | 2 | 2 | 2 | - | - | - | - | - | - | nausea/vo<br>miting<br>(not<br>chemother<br>apy<br>associated<br>) | APM |
| Piritramid | 59 | 36 | 11 | 18 | - | - | - | 22 | - | dsypnea | German<br>Guideline,<br>APM |
| Pregabalin | 102 | 6 | 0 | 6 | - | - | - | - | - | - | - |
| Quetiapine | 52 | 50 | 33 | 17 | - | - | - | 26 | - | delirium,<br>agitation | APM |
| Tapentadol | 1 | - | - | - | - | - | - | - | - | - | - |
| Tilidin | 2 | 2 | - | 2 | - | - | - | - | - | - | - |
| Venlafaxine | 14 | 14 | 14 | - | - | - | - | - | - | - | - |

With decreasing relevance of the groups for symptom control, the total number of application days as well as the off-label/in-label ratio dropped: the group of medium relevant drugs (40; 43%; e.g. pantoprazole, enoxaparin) accounted for 714/2352 (30%) of all application days; of these 255/714 (36%) were off-label. The group with the lowest relevance accounted for only 9% (210) of all application days with a total of 83 (40%) off-label application days.

## Discussion

Little is known about off-label drug use in palliative care in different national and institutional circumstances. This article provides a first detailed evaluation of off-label use on a palliative care unit in Germany. We aimed to get insights into off-label prescribing and therefore critically evaluated not only the indication but also other areas of off-label use, such as drug dose, dosing interval, route and mode of administration. To our knowledge such an extensive assessment of off-label drug use has not been published for adult palliative care previously.

Off-label use was common in this unit with more than half of all drug uses outside the marketing authorisation. Compared to other studies, the frequency of off-label use was rather high in our sample^1^. This can partially be explained by the fact that we evaluated off-label use not only with regard to indication but to six other aspects of the approval (i.e. dose, mode of application, etc.). The large proportion of off-label use regarding the indications was hardly surprising. Many drugs are not approved for the relatively specific indications in palliative medicine. Still, compared to publications from the UK (14.5%^2^) and the US (up to 35%^18^), the frequency of off-label use was remarkably high. The extent to which the use deviated from the actual approval was very heterogeneous. Midazolam, for example, is authorized for sedation in intensive care but not in palliative medicine. In comparison, the established palliative care indication of dyspnea for opioids is not even close to being covered by the approval in Germany, although very well covered by scientific evidence^19, 20^. In our study, the application mode was the second most common cause for off-label use. The reason for this was almost exclusively the non-approved mixing of different substances in a syringe and the subsequent application as continuous infusion. Since data on the compatibility of the mixtures used and on the benefit of a continuous infusion compared to intermittent injections are only available to a very limited extent, this practice must be critically questioned and discussed in more detail. A route of administration beyond the approval only came in fourth place in our evaluation. However, when considering subcutaneous administration alone, our assumptions of more common off-label use were confirmed. These findings are consistent with other studies evaluating subcutaneous adminstration in palliative care (62-85%^2, 21^).

Among drugs and drug groups frequently used off-label were ketamine, antipsychotics, antidepressants, drugs for gastrointestinal symptoms, corticosteroids and opioids. These were frequently identified in other studies in the context of off-label use and palliative care aswell^2, 18, 22^. Based on these findings it seems that common symptoms in advanced disease like dyspnoea, nausea or vomiting either lack authorized therapies or licensed alternatives are not used. Possible reasons for the latter, however, cannot be determined due to the retrospective study design.

An important finding is the high number of overall off-label application days, the proportion of off-label days in the group of “highly relevant” palliative care drugs, and the decreasing number of overall and off-label application days with decreasing relevance of a drug. This results underpins the high relevance of off-label use in palliative medicine. However, this does not allow for conclusions on the justification of off-label drug use in palliative medicine. Especially the lack of evidence to support off-label use and the high proportion of off-label days without evidence need to be noted. This proportion is rather high compared to other studies (3-11%^2, 18^). Although we have compared our data with two well-known and established references^14, 23^, this approach does only allow limited conclusions on the quality of the underlying evidence.

We assessed off-label use with regard to every aspect of the marketing authorisation, the relevance of drugs for palliative care and the availability of explicit literature references. Data available so far still only allows limited insights into off-label prescribing behaviour in different institutions and in different national contexts, though. Reasons that led to a therapeutic decision cannot be illustrated. It is also not clear what role patient-specific or infrastructural factors played in the treatment selection. To be able to continue working with our and previous findings, it is important to have the opportunity to compare off-label use in different institutions and different countries. Such data can serve as a benchmark for quality assurance; they can help to identify areas that seem to represent therapy standards but with no or very little available evidence on the other. They can also be used to identify therapeutic strategies that may be outdated by other approaches. Based on such comparisons it would also be possible to develop quality indicators for off-label use in palliative care. Our results demonstrate that there are multiple opportunities for pharmacists to support off-label use of drugs in clinical routine, especially when evaluating a treatment or possible treatment alternatives.

This study has several limitations. Data extraction was restricted to one month in one palliative care unit. Therefore, data collection on off-label use from different palliative care institutions in Germany is currently being prepared as a follow-up project. Retrospective data collection relies on the quality of the documentation as wrong or missing data cannot be replaced or completed. We were able to extract the required data for all patients that were treated during the period analysed and, in most cases, ambiguities could be resolved based on the patient records. The data presented here is from 2017 and might be considered outdated. However, for a retrospective evaluation of prescribing data, a time delay until the data is available should always be expected. Furthermore an update and extension of the German Guideline on Palliative Medicine for Patients with Incurable Cancer^24^ has been published and its influence on prescribing behaviour is not clear. It will therefore be interesting to compare whether it had an impact on off-label prescribing behaviour.

Off label use is an integral part of palliative medicine. With very strict consideration, which does not only focus on the indication, the extent may be even greater than previously known. Off-label use should not be a routine but rather encourage to question a therapy. At the same time, however, it must be possible to prescribe well-established and scientifically sound therapies without justification if appropriate for an individual patient. Ultimately, it is important to critically question the necessity of every drug - whether inside or outside the marketing authorisation, for every patient. As an important first step, the data presented here allow for a more precise characterization of various aspects of off-label use in everyday clinical practice in order to use this knowledge to derive concrete further measures for research and clinical practice. As a follow-up to this pilot study, the next step will be to collect and evaluate data on off-label use in various palliative care settings. These results will then be used, in part, for quality assurance in the off-label drug use in palliative care.

## Data Availability

Data referred to in the manuscript is available from the corresponding author upon reasonable request

## Acknowledgements

The authors would like to thank Stefanie Büsel for assisting in the data extraction process.

